# Unveiling Vulnerabilities in Maternal-Child Health amidst COVID-19: A Cross-Sectional Study on Food Insecurity and Socio-Demographic Disparities in the U.S.

**DOI:** 10.1101/2023.11.28.23299080

**Authors:** Zoe Henkes, Maria J. Romo-Palafox

## Abstract

**Objective:** Early infancy is a critical time of development when stresses, nutritional deficiencies, and other challenges have lifelong consequences. Social distancing regulations due to the COVID-19 pandemic in 2020 led to abrupt changes in work status, childcare accessibility, and food availability.

**Design:** This cross-sectional study assessed responses regarding experiences during the COVID-19 pandemic, including food insecurity (validated two-question screener), WIC use, and changes in childcare accessibility and work status. Data were assessed using logistic regressions while controlling for sociodemographic factors.

**Setting:** National U.S. online survey in July–August 2020.

**Participants:** U.S. mothers (n=1861) with infants ≤12 months old.

**Results:** We detected a 34% increase in perceived food insecurity during July–August 2020 compared to that before the pandemic. Hispanic mothers had 74% higher odds of becoming food insecure than non-Hispanic White mothers. Mothers with infants <9 weeks old had a 7% increase in WIC utilization, although no overall increase in WIC usage was detected. Most mothers (71%) reported moderate or extreme impacts from the pandemic, with higher odds associated with childcare interruptions, working from home, and identifying as Hispanic or non-Hispanic Black.

**Conclusions:** Our findings reveal specific sociodemographic groups of mothers with infants who were especially vulnerable during the COVID-19 pandemic. These insights hold significant value for tailoring supportive programs, equipping these groups for potential socioeconomic upheavals, and aiding their transition into the post-pandemic world.

## Introduction

The first 1,000 days of an infant’s life, from gestation to two years old, are critical for growth, neurological development, cognition, and learning ^(1)^. Infants must receive adequate nutrition and have a stable care environment during this time. Toxic stress and economic instability that affects caregivers and infants can be detrimental and enduring ^(2)^. Early childhood malnutrition can result in lower IQ scores, difficulties in school, and behavioral problems ^(2,3)^. The social distancing and shelter-in-place orders during the COVID-19 pandemic changed mothers’ work and income status, childcare availability, and food security. As a result, infants may have experienced significant challenges during their first 1,000 days, which could have lasting effects on their health and well-being.

In the United States, about 15% of households with children under the age of six experienced food insecurity in 2020 ^(4)^, with Black, Hispanic, and other communities of color disproportionately represented ^(4)^. Although impoverished households are typically at greatest risk for becoming food insecure, sudden life changes such as divorce or loss of employment can lead families to become food insecure even if they were not originally low-income ^(2)^. On a global scale, the COVID-19 contingency induced new stresses and social changes, and experts estimated in late May 2020 that approximately 86 million children in low- and middle-income countries had a high risk of sliding into poverty by the end of the calendar year ^(5)^. These mass changes in household income and food security status caused by the COVID-19 pandemic could have lasting effects on the health of US infants during their first 1,000 days.

Women have historically taken on a higher proportion of domestic childcare responsibilities than men, and that gap continued to widen during the COVID-19 pandemic. In 2018, women in the US were responsible for 1.5 times the amount of unpaid care work (primarily household and childcare responsibilities) than men ^(6)^. In the spring of 2020, childcare center closures forced many women to take on even further increases in childcare responsibilities ^(7–9)^.

The work status of many women changed during the spring and summer of 2020 due to unemployment, furlough, transitioning to remote work, or the need for alternative childcare in light of widespread closures of childcare facilities. Income status affected pandemic-induced outcomes, with 17% of low-income US women quitting work due to the pandemic, whereas only 5% of high-income women had to quit work ^(10)^. Other factors impacting this stratification included field of work, flexibility of hours, salaried versus hourly pay, and the ability to work remotely ^(8)^. However, the loss of childcare is a distinct factor that distinguishes the COVID-19 pandemic from prior episodes of economic recession resulting in higher unemployment rates among mothers of young children ^(7,10,11)^. The magnitudes of these socioeconomic changes are concerning because disrupted childcare and education services impacted more than 40 million children worldwide as of July 2020 ^(5)^.

The objective of this study was to quantify food insecurity, Special Supplemental Nutrition Program for Women, Infants, and Children (WIC) use, and changes in work and childcare experienced by US women with children under 12 months of age in July–August 2020. We also identified sociodemographic factors associated with the risk of becoming food insecure, and women’s perceived impact of the COVID pandemic on their daily lives. This research expands existing evidence of gender and racial inequalities in work and income during natural disasters or economic downturns. These results also inform advocates and policymakers seeking to improve maternal leave policies, increase childcare access and support, and promote proper nutrition for mothers and young children.

## Methods

### Survey and participants

Centiment is a national online survey panel that recruited a sizeable sample of US mothers in July–August 2020 (*n*=1861). The platform provides redeemable points to participants for completing voluntary surveys and does not offer direct monetary incentives. The survey items were pretested using a convenience sample of 10 mothers of infants younger than 12 months of age. Subjects evaluated questions for clarity, interpretation, and recall issues during one-on-one cognitive interviews.

This cross-sectional study utilized data from the Centiment online survey of US mothers (*n*=1861) exploring impacts of the COVID-19 pandemic and stay-at-home orders on women’s work status, childcare accessibility, food insecurity, and breastfeeding experiences. Participants include women who self-identified with children aged 0–12 months.

### Demographic characteristics

Mothers reported their age, highest education level, race, ethnicity, and annual household income. Education level was categorized into three groups: less than high school, high school diploma, or GED; some college or 2-year college; 4-year college or higher. Race and ethnicity were categorized as non-Hispanic White, non-Hispanic Black, Hispanic, Asian, American Indian, Pacific Islander, or mixed/other. Our researchers combined Asian, Pacific Islander, American Native, and mixed/other into one group for statistical power. The age of the women’s youngest child was categorized as <9 weeks old, 10–26 weeks old, or 26–52 weeks old.

### Impacts of the COVID-19 pandemic on survey respondents and childcare accessibility

Survey respondents reported details about how the shelter-in-place orders affected the amount of their time spent at home. The impact of the COVID-19 pandemic on daily life was rated using a 5-point Likert scale (from “not at all impacted” to “extremely impacted”). Respondents selected specific pandemic-related challenges from among 18 potential challenges, including changes in health behaviors, physical and mental health challenges related to COVID-19 disease, and impacts of the COVID-19 contingency.

### Food insecurity screening and WIC use

Food insecurity was determined using the 2-question food insecurity screening tool developed by Hager et al ^(12–14)^. Survey respondents were asked to recall the time before COVID-19 pandemic contingencies, including the closure of schools, daycare, and educational institutions in their respective localities. Respondents provided information about their food security before the pandemic and during July–August 2020; specifically, whether (a) they were worried whether food would run out before they got money to buy more food, or (b) whether the purchased food did not last long enough and they did not have money to buy more food. Per the tool’s methodology, we coded “often true” and “sometimes true” responses to either statement as positive for food insecurity. Respondents also reported their use of the WIC program before the pandemic and during July–August 2020, and we analyzed pandemic-induced changes in program utilization.

### Pandemic-induced changes in childcare and work status

Survey respondents reported changes in childcare accessibility based on a list of five options ranging from “never used/didn’t plan to use childcare,” to “alternative childcare,” to “stop using any form of childcare.” Work status changes were assessed based on a list of 12 options, including “continued to go to work as normal,” “homemaker, the situation has not changed,” “working from home,” “taking parental leave,” “staying at home, unable to work,” and “currently unemployed.”

### Statistical analysis

We analyzed descriptive statistics for sociodemographics, food insecurity, WIC use, perception of time spent at home, perception of COVID-19 impacts, childcare, and work status. One logistic regression model computed the odds of food secure mothers becoming food insecure using sociodemographics as predictors. Then, two logistic regression models computed the odds of mothers perceiving either a moderate or extreme, grouped together and referred to as “moderate-to-extreme” or only extreme disruption to their daily lives using sociodemographics, childcare, and work status as predictors. All data analyses were performed in 2021 using SAS version 9.4 (SAS Institute, Inc.).

## Results

The final survey sample included 1,861 US women who were predominately aged 18–35 years. Most mothers had infants aged 26–52 weeks old (43%), whereas 31% had infants aged 10–26 weeks old, and 25% had infants aged <9 weeks old. This survey sample had more respondents (43%) with an annual income less than $40,000 (low-income group) than those in middle- and high-income groups (Table 1). The education levels of mothers were evenly distributed. Chi-square tests did not detect notable trends or differences in race or ethnicity (58% non-Hispanic White), level of education, or household income between child age groups (Table 1). Survey respondents with children aged 26–52 weeks were significantly older (68% were >26 years old, *p*<0.0001).

**Table 1.** Sociodemographic characteristics and percent time spent at home by mothers of children under 12 months when stay-at-home orders for COVID-19 contingency began (*N*=1861, July–August 2020)

|  | Child's age |  |  |  |  |  |  |  |
| --- | --- | --- | --- | --- | --- | --- | --- | --- |
|  | <9 weeks |  | 10–26 weeks |  | 26–52 weeks |  | Total |  |
|  | *n* | % | *n* | % | *n* | % | *n* | % |
| Sample | 469 | 100 | 583 | 100 | 809 | 100 | 1861 | 100 |
| Mother's race/ethnicity |  | 25 |  | 31 |  | 43 |  |  |
| White non-Hispanic | 264 | 56 | 329 | 56 | 490 | 61 | 1083 | 58 |
| Black non-Hispanic | 72 | 15 | 80 | 14 | 113 | 14 | 265 | 14 |
| Hispanic | 72 | 15 | 108 | 19 | 118 | 15 | 298 | 16 |
| Asian | 17 | 4 | 23 | 4 | 30 | 4 | 70 | 4 |
| American Indian | 6 | 1 | 6 | 1 | 4 | 0 | 16 | 1 |
| Pacific Islander | 3 | 1 | 3 | 1 | 2 | 0 | 8 | 0 |
| Mixed or other | 35 | 7 | 34 | 6 | 52 | 6 | 121 | 7 |
| Mother's age |  |  |  |  |  |  |  |  |
| 18–25 | 213 | 45 | 241 | 41 | 256 | 32 | 710 | 38 |
| 26–35 | 233 | 50 | 292 | 50 | 454 | 56 | 979 | 53 |
| >35 | 23 | 5 | 50 | 9 | 99 | 12 | 172 | 9 |
| Mother's education |  |  |  |  |  |  |  |  |
| Less than high school, high school, or GED | 153 | 33 | 184 | 32 | 237 | 29 | 574 | 31 |
| Some college, 2-year college | 157 | 33 | 195 | 33 | 295 | 36 | 647 | 35 |
| 4-year college or higher | 159 | 34 | 204 | 35 | 277 | 34 | 640 | 34 |
| Household income |  |  |  |  |  |  |  |  |
| <\$40,000 | 204 | 43 | 253 | 43 | 340 | 42 | 797 | 43 |
| \$40,000–\$75,000 | 103 | 22 | 148 | 25 | 216 | 27 | 467 | 25 |
| >\$75,000 | 122 | 26 | 151 | 26 | 207 | 26 | 480 | 26 |
| Prefer not to say | 40 | 9 | 31 | 5 | 46 | 6 | 117 | 6 |
| Percent of time spent at home |  |  |  |  |  |  |  |  |
| >75% | 291 | 62 | 362 | 62 | 508 | 63 | 1161 | 62 |
| 50–75% | 101 | 22 | 113 | 19 | 174 | 22 | 388 | 21 |
| 25–50% | 53 | 11 | 82 | 14 | 96 | 12 | 231 | 12 |
| ≤25% | 24 | 5 | 26 | 4 | 31 | 4 | 81 | 4 |

### Perceived impacts and challenges of the COVID-19 pandemic

Most mothers (71%) perceived that the COVID-19 pandemic significantly impacted their day-to-day lives: 36% of survey respondents reported extreme impacts and 35% reported moderate impacts (Table 2). Only 2% reported no impacts. A chi square test of these data indicated that 41% of mothers with children aged <9 weeks were extremely impacted, whereas 32% of those with children aged 26–52 weeks were extremely impacted (*p*<0.01).

**Table 2.**
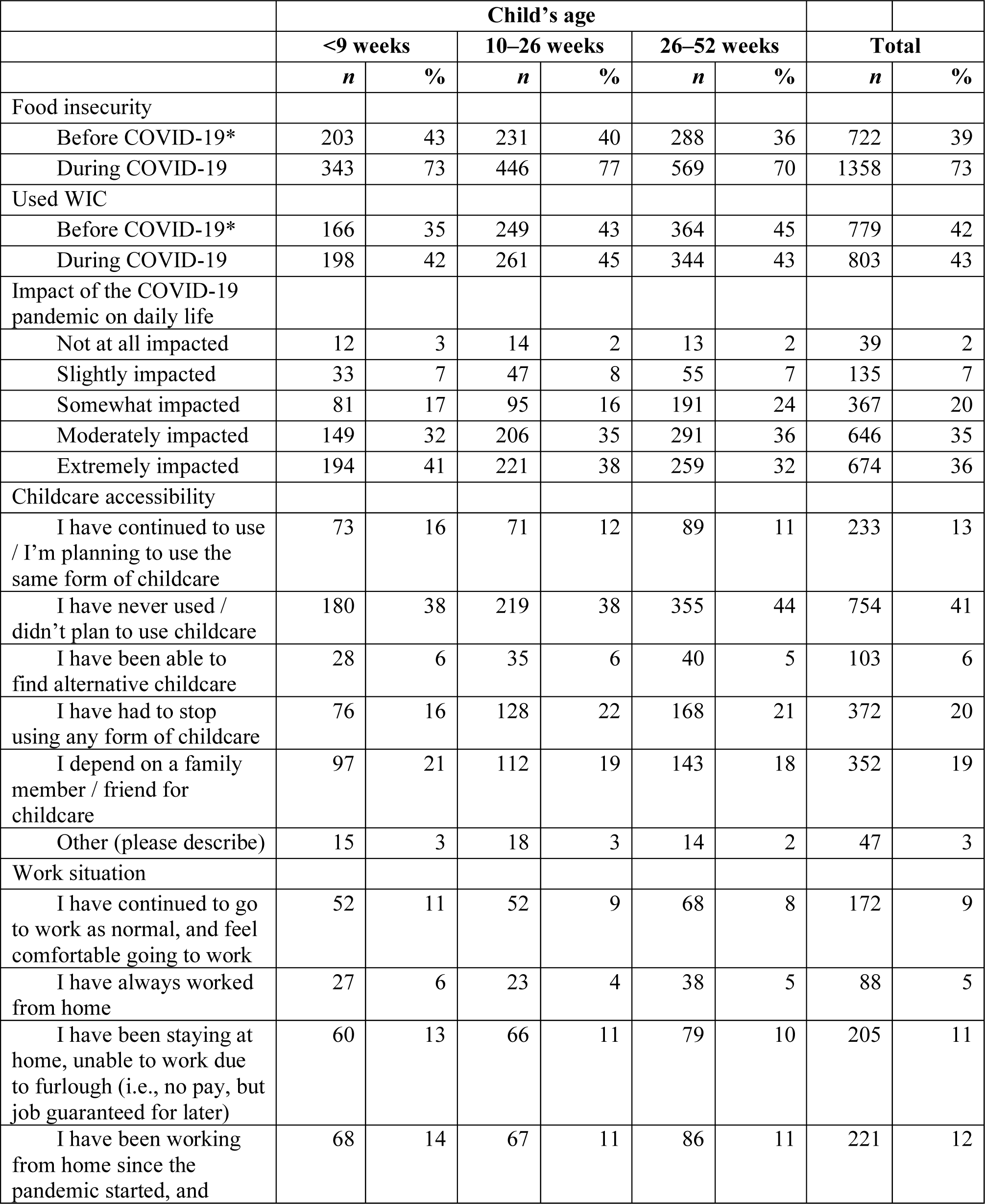
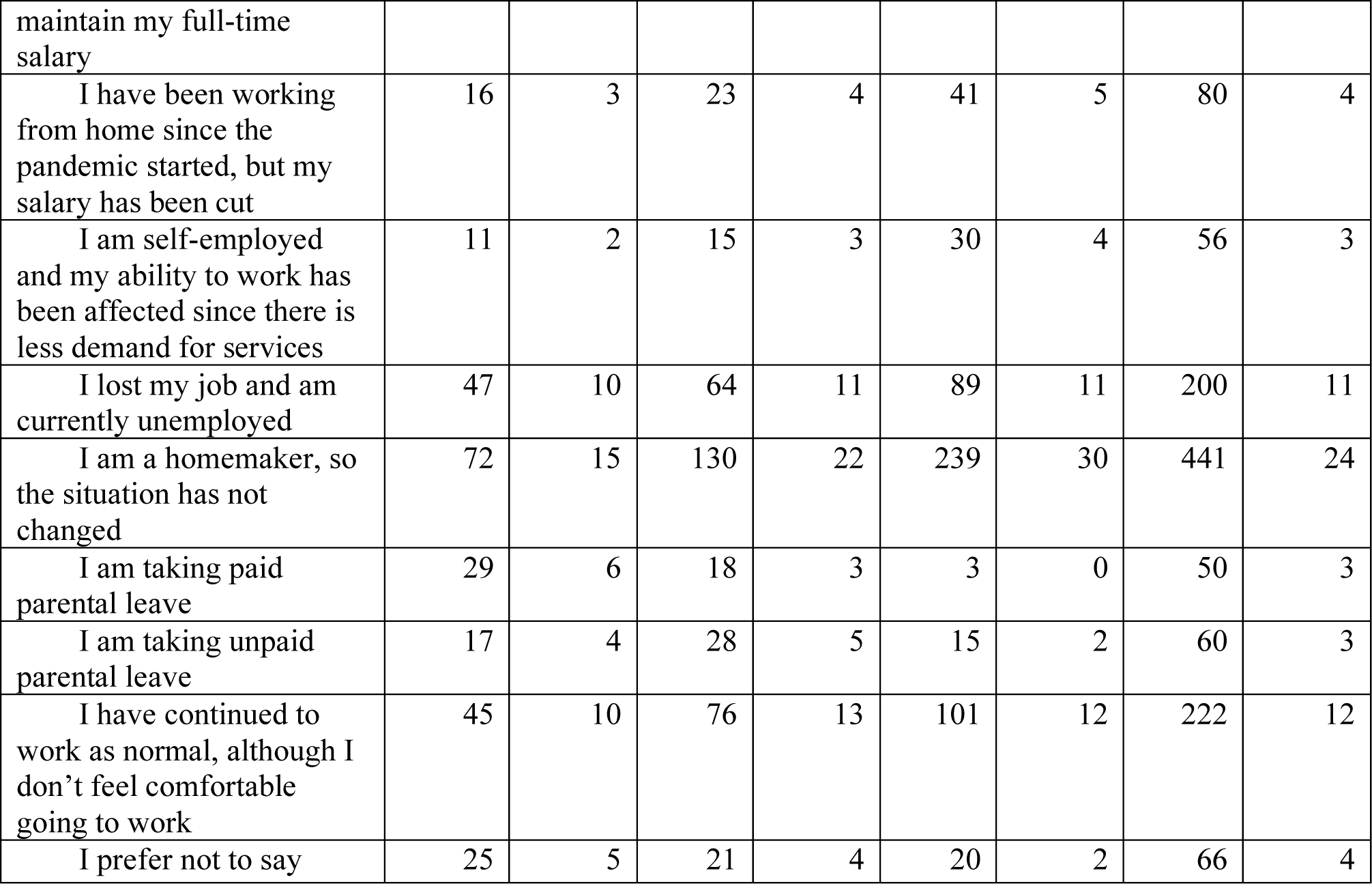
Mothers’ perceptions of the effects of stay-at-home orders on food security, daily life, work status, and childcare accessibility during the early COVID-19 contingency (*N*=1861, July–August 2020)

Survey respondents selected specific challenges relevant to their situation from among a list of 18 potential challenges caused by the COVID-19 crisis. More than 60% of respondents selected “worrying about friends, family, partners, etc.” More than 50% of respondents selected “frustration or boredom,” “fear of getting COVID-19,” and “more anxiety,” respectively. Respondents had significant anxiety regarding contracting (60%) and/or spreading (27%) COVID-19, although <10% selected “being diagnosed with COVID-19.” Other mental health challenges included changes in sleep (44%), loneliness (39%), and increased depression (37%). Approximately 25% of respondents reported that deficiencies in basic supplies (e.g., food, water, medication) were pandemic-specific challenges.

### Sociodemographic factors and perceived pandemic impact

After controlling for sociodemographic characteristics, work status and conditions, and childcare accessibility, Hispanic mothers had double the odds (OR=2.0, 1.4-2.9) of selecting “moderately” or “extremely” impacted by COVID-19 compared to non-Hispanic White mothers (Fig. 2). However, Hispanic race/ethnicity was not significant when analyzing only for “extreme” impacts, and non-Hispanic Black mothers had 1.5 times higher odds of selecting “extremely” impacted (OR=1.5, 1.1–2.0) (Fig. 3). There was no effect of education level or household income on the perceived impact of COVID-19.

**Fig. 1.**
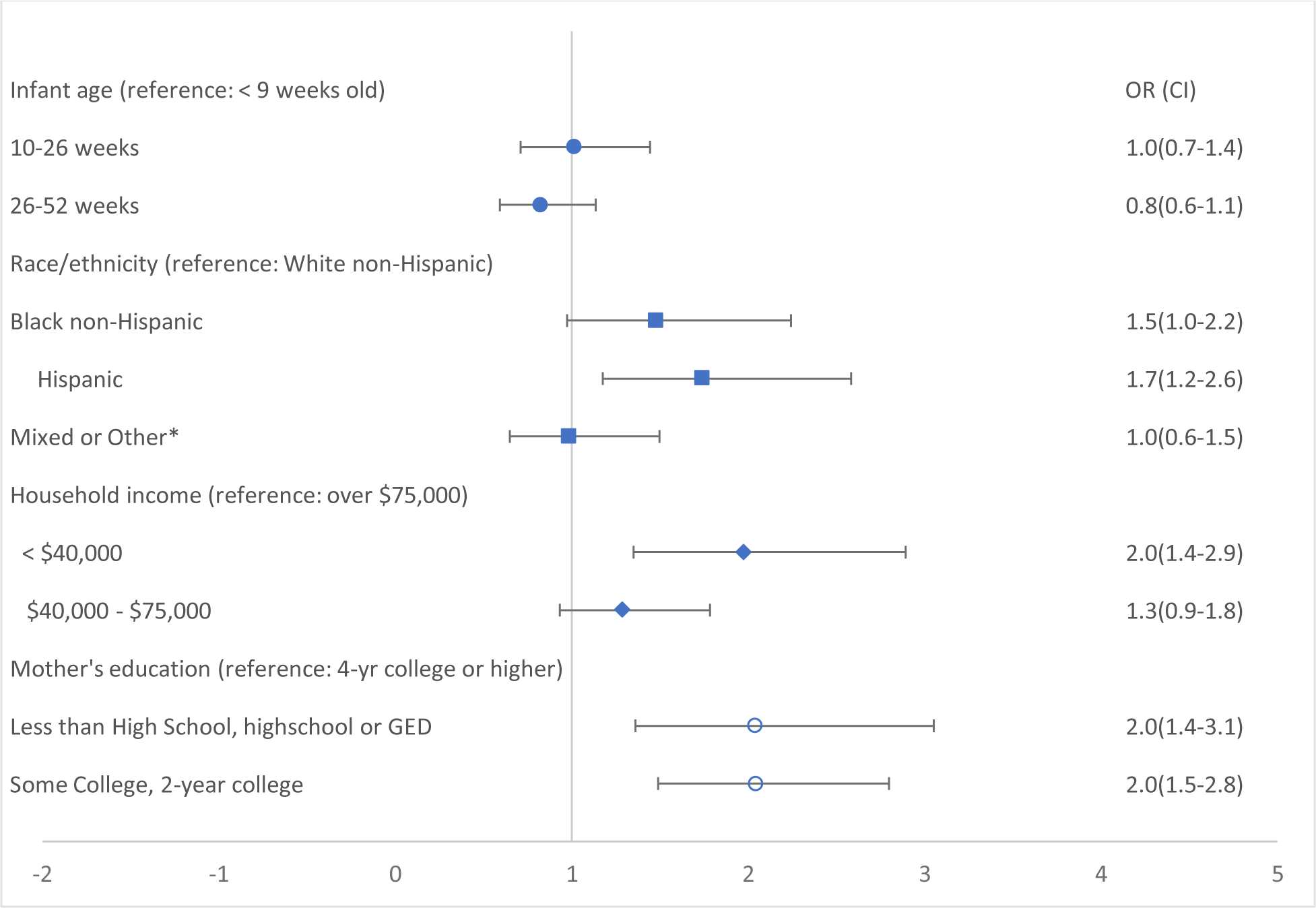
Odds of food secure mothers becoming food insecure by sample characteristics (*n*=1069, 2020)

**Fig. 2.**
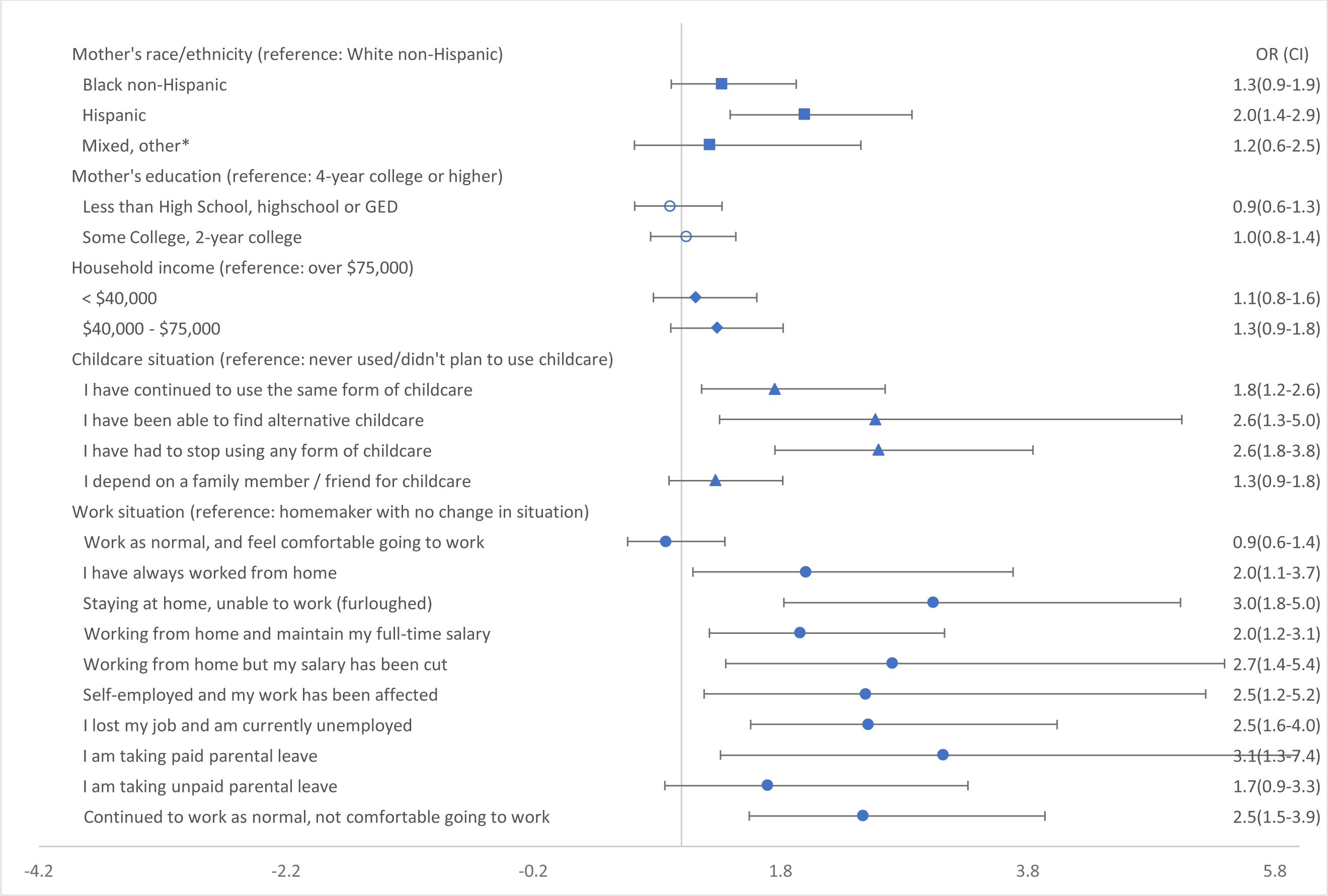
Odds of mothers reporting moderate-to-extreme impacts of the COVID-19 pandemic on daily life by demographics, childcare accessibility, and work status (July–August 2020, *n*=1,473)

**Fig. 3.**
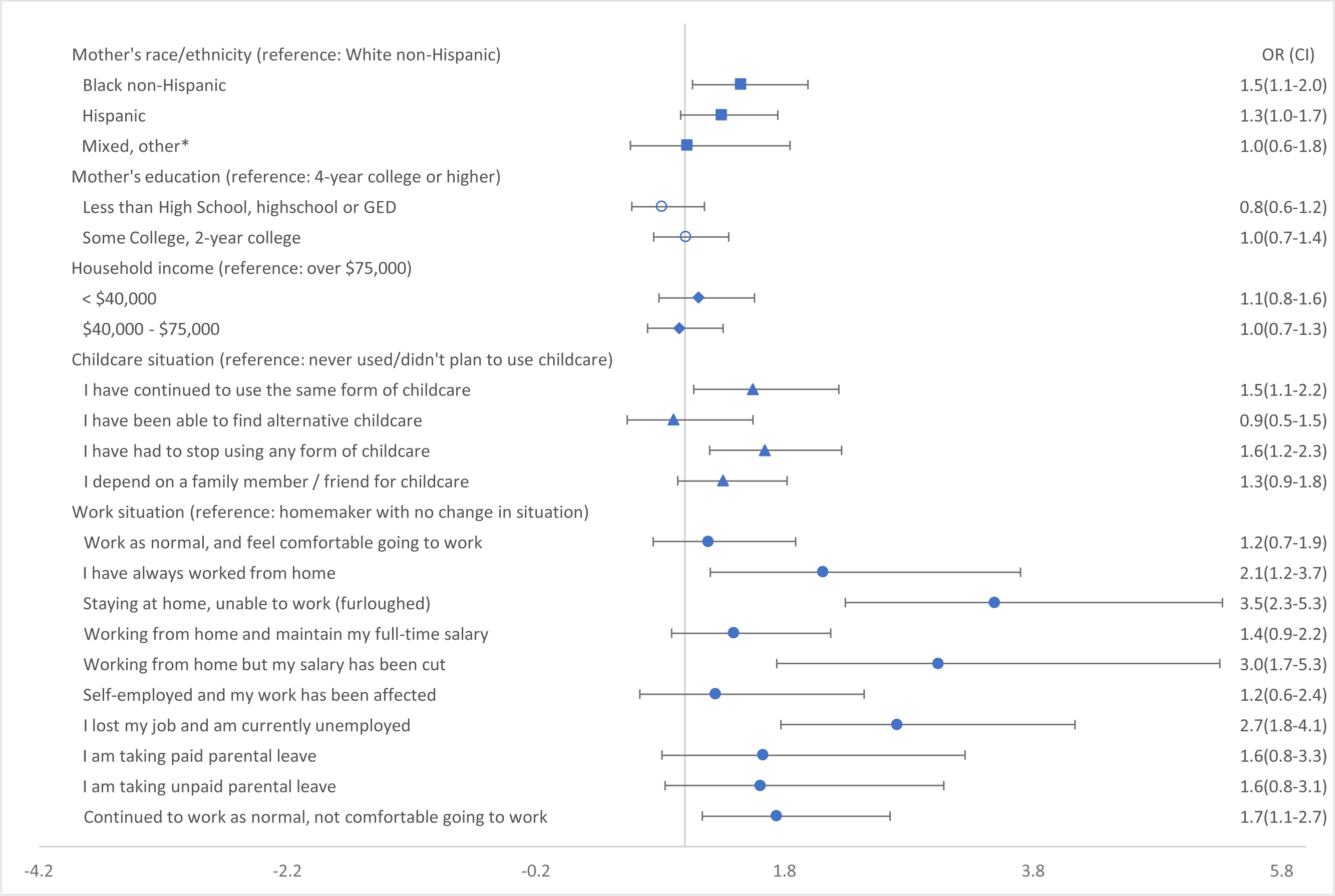
Odds of mothers reporting extreme impacts of the COVID-19 pandemic on daily life by demographics, childcare accessibility, and work status (July–August 2020, *n*=1,473)

### Changes in food security, WIC use, and risk of becoming food insecure

Responses to the validated two-question food insecurity screening tool indicated that 39% of respondents experienced food insecurity before the pandemic, with a 34% increase in food insecurity perception during July–August 2020. WIC use did not significantly differ before the pandemic (42%) and during in July–August 2020 (43%); however, mothers of younger infants (<9 weeks old) reported a 7% increase in WIC use, whereas mothers of older children (10–52 weeks) averaged a 0.4% decrease (Table 2).

After controlling for sociodemographic characteristics, Hispanic mothers had 74% higher odds of becoming food insecure during the early pandemic (July–August 2020) than non-Hispanic White mothers. Survey respondents with an annual household income less than $40,000 had double the odds (OR=2.0, 1.4-2.9) of becoming food insecure than those with an annual income greater than $75,000. Education levels of below high school, high school, or GED (OR=2.0, 1.4-3.1) and some or 2-year college degree (OR1.5-2.8) were associated increased odds of becoming food insecure compared to those with a 4-year college degree or higher. The infant’s age, 10-26 weeks (OR=1.0, 0.7-1.4) or 26-52 weeks (OR=0.8, 0.6-1.1) did not affect the odds of becoming food insecure compared to infants <9 weeks old(Fig. 1).

### Childcare accessibility and perceived pandemic impact

At the time of the survey (July–August 2020), 41% of mothers reported they had never used nor planned to use any form of childcare, 13% planned to continue using the same form of childcare during the COVID-19 contingency, 6% found alternative childcare, and 20% had to stop using childcare during the early pandemic.

Childcare accessibility significantly affected the perceived impact of the COVID-19 crisis on daily life. Those who never used or did not plan to use childcare were unaffected, whereas all other childcare situations were associated with increased odds of perceiving a moderate-to-extreme impact of the pandemic. Those who had to stop using any form of childcare during the pandemic were associated with the highest odds of perceiving a moderate impact of COVID-19 (OR=2.6) compared to those who did not use any form of childcare (Fig. 3). The effects of being “able to find alternative childcare” (OR=0.9, 0.5-1.5) and “depending on a family member/friend for childcare” (OR=1.3, 0.9-1.8) were not significant when considering only extreme impact. The effects on those who “continued to use the same form of childcare” and “had to stop using any form of childcare” were higher for moderate-to-extreme impacts (OR=1.8, 1.2-2.6 and OR=2.6, 1.8-3.8, respectively) than for extreme impacts (OR=1.5, 1.1-1.2 and OR=1.6, 1.2-2.3, respectively).

### COVID-19 pandemic impact on time spent at home and work status

Most mothers (62%) reported spending more than 75% of their time at home during July–August 2020, whereas only 4% of survey respondents spent ≤25% of their time at home. Approximately 25% of mothers were homemakers with no change in their work status during the pandemic (30% had children aged 26–52 weeks and 15% had children aged <9 weeks). At the time of the survey (July–August 2020), 9% of respondents continued to work as usual and were comfortable doing so, and 12% continued to work but did not feel comfortable. A total of 22% of respondents lost their job or were furloughed during the early months of the COVID-19 contingency.

Work status was a significant predictor of women’s perceived impacts of the pandemic when controlling for other factors using homemakers with no change in their work status as the reference group. Mothers who always worked from home had two times higher odds (OR=2.0, 1.1-3.7) of perceiving moderate-to-extreme impacts of the pandemic than homemakers who had no change in their work status (Fig. 3). Mothers who were furloughed and then stayed home had three times the odds (OR=3.0, 1.8-5.0) of perceiving moderate-to-extreme impacts. Those who began to work from home during the pandemic and maintained full salary (OR=2, 1.2-3.1) or experienced a pay decrease (OR=2.7, 1.4-5.4) had higher odds of reporting moderate-to-extreme impacts. Survey respondents who were self-employed and had their workload affected by the pandemic had 2.5 times the odds (OR=2.5, 1.2-5.2) of reporting at least a moderate impact compared to homemakers who had no change in their work status. Respondents who lost their job and were unemployed at the time of the survey had 2.5 times higher odds (OR=2.5, 1.6-4.0) of reporting at least a moderate impact. Mothers who were taking a paid parental leave had 3.1 times higher odds (OR=3.1, 1.3-7.4) of perceiving moderate-to-extreme impacts compared to homemakers, whereas those taking an unpaid parental leave did not demonstrate significantly increased odds of perceived impact (OR=1.7, 0.9-3.3). Those who continued to work outside of the home but were not comfortable going to work had 2.5 times higher odds (OR=2.5, 1.5-2.9) of reporting moderate-to-extreme impacts of the pandemic than homemakers with no change in their work status.

Considering all work status groups, those who were furloughed were most likely to report extreme impacts of the pandemic (OR=3.5, 2.3-5.3) with a slightly higher strength of association compared to those perceiving moderate-to-extreme odds (OR=3.0, 1.8-5.0) (Fig. 3). Those who always worked from home had 2.1 times higher odds (OR=2.1, 1.2-3.7) of reporting extreme impacts than the reference group of homemakers with no change in work status. Survey respondents who worked from home with reduced salary since the start of the pandemic had three times higher odds (OR=3.0, 1.7-5.3) of reporting extreme impacts of the pandemic. Those who lost their jobs had 2.7 times higher odds (OR= 2.7, 1.8-4.1) of reporting extreme impacts of the pandemic. Respondents who worked from home with no change in salary did not have significantly higher odds (OR=1.4, 0.9-2.2) of reporting extreme impacts of the pandemic. Those who continued to work outside the home but felt uncomfortable going to work reported extreme impacts of the pandemic (OR=1.7, 1.1-2.7) compared to homemakers with no change in work status, although the strength of association was lower than that of the moderate-to-extreme impact group (OR=2.5, 1.5-3.9).

## Discussion

Our results indicated that mothers with infants aged 0–12 months had higher food insecurity rates during the COVID-19 pandemic, highlighting the negative impacts on health and financial status during a global pandemic. Although food insecurity is not a direct measure of nutritional status, it does reflect a higher risk for inadequate nutrition ^(15)^ and obesity ^(16,17)^. National food insecurity rates have been declining in recent years, with 10.5% of US households qualifying as food insecure in 2019 ^(4)^. The 2020 USDA annual household food security study reported that overall household food insecurity rates did not change from 2019 to 2020 ^(4)^. However, food insecurity and very low food security increased significantly from 2019 to 2020 for specific subgroups such as households with children (13.6% to 14.8%, respectively) ^(4)^. Our analyses detected higher baseline rates (before the pandemic) of food insecurity than those observed in national data ^(4)^. However, the high rates of survey respondents who retrospectively perceived food insecurity before the pandemic could be due to our sample sociodemographics (primarily lower income mothers of infants) as this population was already at risk of higher rates of food insecurity. Households categorized as low income (28.6%), non-Hispanic Black (21.7%), Hispanic (17.2%), and single mothers (27.7%) all had significantly higher rates of food insecurity than the national average (10.5%) in 2020^(4)^. Other US studies assessing changes in food insecurity due to the pandemic also reported higher initial food insecurity rates than national data for adults ^(18)^ and households with children ^(19–21)^. One explanation for this is that retrospective perceptions may be influenced by current situations. Studies performed in spring and summer of 2020 reported similar increases in food insecurity during the pandemic, indicating that the pandemic greatly affected access to food ^(18,19,22)^.

Our results detected specific sample parameters that impacted the odds of becoming food insecure. A previous study conducted in Los Angeles County, California, reported that Hispanic families experienced higher rates of food insecurity during the COVID-19 pandemic than non-Hispanic White households ^(23)^, consistent with our results from the current national sample. However, our results detected significant associations between Hispanics and food insecurity when controlling for income, unemployment, and age, which was not observed in the previous study ^(23)^. This suggests that mothers with young children could be more vulnerable to becoming food insecure than the general population. Several studies reported associations between educational attainment and food insecurity in relationship to obesity rates, but no studies specifically assessed the effect of education level on food insecurity ^(24,25)^. The US Department of Agriculture reported a positive correlation between low-income households and food insecurity ^(4)^.

Although food insecurity increased during the COVID-19 pandemic, we did not detect a corresponding increase in WIC usage, consistent with the reports of other studies ^(20)^. Families have historically cited many barriers to utilizing WIC services, and these barriers appear to have been exacerbated during the pandemic. These barriers include stigma from retailers, limited product choices, and limited product supplies due to stockpiling. These challenges resulted in providers traveling longer distances to stores, visiting multiple stores, and shopping more often because food didn’t last as long, which led to increased feelings of anxiety and stress surrounding COVID-19 ^(26)^. Other barriers cited by Hispanic-American immigrant families included repercussion fears, logistical barriers, misinformation or lack of information, and lack of perceived need ^(27)^. Niles et al. reported that those who became food insecure during the pandemic were less likely to utilize government food assistance programs such as WIC compared to those who were food insecure before the pandemic ^(19)^. These results indicate a lack of resources and knowledge for those with newly induced food insecurity. Given the established link between food insecurity and adverse health outcomes for infants, attention should be paid to vulnerable populations at higher risk of becoming food insecure during national crises.

One health system in Los Angeles county reported a 24% increase in WIC usage from February to June 2020 ^(21)^ by leveraging virtual program enrollment and rapidly expanding online services. The state of California experienced a 14% increase in WIC participation during the early pandemic, but the study did not assess WIC usage in other states ^(21)^. This suggests that WIC usage may be highly variable across health systems, and federal measures should be implemented to increase WIC accessibility across the US in times of economic downturn.

New mothers (infants <9 weeks old) had higher WIC usage during the pandemic than before, whereas mothers with older infants (10–52 weeks) did not increase their utilization of WIC. The reasons for this trend are unclear, but suggest that new mothers have greater awareness of resources such as WIC than mothers with older infants, or that those who did not need WIC before the pandemic may lack knowledge about available resources.

After controlling for other sociodemographic factors, childcare accessibility, and changes in work status, non-Hispanic Black and Hispanic mothers were more likely to report higher levels of impact due to the pandemic than non-Hispanic White mothers. Several studies reported higher rates of COVID-19 incidence and death among communities of color ^(28–32)^. These results highlight racial and ethnic disparities impacting health outcomes in the US. Structural racism (in the form of residential segregation, incarceration, poorer educational attainment, lower economic indicators, and higher unemployment) is a strong contributor to these striking disparities ^(28,30,33)^. Racial segregation leads to crowded living and public transportation conditions and food deserts, thereby impacting the ability to socially distance and obtain healthy food, which increases pandemic-induced stress and anxiety. Communities of color historically have higher burdens of chronic diseases such diabetes, chronic obstructive pulmonary disease, hypertension, and ischemic heart disease, all of which were identified as common comorbidities in COVID-19 deaths ^(32)^. Our survey did not measure COVID-19 cases or death rates, but being surrounded by community and family members affected by COVID-19 would be anticipated to impact the respondents’ experiences of the pandemic.

Although mothers of Hispanic, Black, and other communities of color were affected by shared racial disparities, Hispanic mothers perceived that the pandemic had moderate-to-extreme impacts on their daily lives (but not extreme impacts), whereas Black mothers experienced the opposite effect. Several studies considered the “Hispanic paradox,” where Hispanics have better health outcomes than predicted. One of these studies assessed whether the Spanish language affected stress perceptions, and reported that wider expression of emotions through language enhanced optimism and modulated stress responses ^(34)^. Another study suggested that culture-specific resiliency factors impact health outcomes; solidarity, shared core values, and shared experiences of Hispanic community members may enhance resilience in the face of adversity ^(35)^.

Organizing childcare is a significant consideration for many families that impacts work, financial, and lifestyle decisions. Changes in childcare accessibility during stay-at-home orders greatly disrupted daily life. Women continued to provide higher proportions of childcare on top of work responsibilities during the pandemic ^(7–9)^. The closure of childcare facilities led to children staying home and increased the responsibilities of mothers. Many childcare facilities that were initially temporarily closed during the early pandemic have become permanently closed due to long-term economic factors and/or low staffing/enrollment after reopening. A statewide survey of licensed childcare providers in Colorado indicated that 9.4% of providers closed during early stay-at-home orders, 20% of those facilities may not reopen, and 6% were closed permanently ^(36)^. This will have long-term effects as families seek new facilities or transition to full-time care of their own children. A recent study reported that 10% of US women quit their jobs due to the pandemic, with 50% citing loss of childcare as a main reason ^(11)^.

Childcare facility closures may disproportionately affect communities of color and expand childcare deserts ^(37)^. Changes in work status significantly affected the perceived impact on daily life during the COVID-19 pandemic. Studies conducted during previous economic downturns reported that changes in work and income leading to economic hardship have negative consequences, particularly on mental health ^(38,39)^. Analyses of an economic recession in southern Europe in 2017 indicated that financial loss and lack of structured time had the strongest associations with decreased life satisfaction. For women, living in social isolation and with an unemployed partner was also associated with lower life satisfaction ^(40)^. Our study did not measure marital status or partner work status, and these factors could have contributed to the survey responses. A study of Swedish workers who transitioned to telework during the pandemic reported reduced workloads, performance, and well-being, which could impact perceptions of the pandemic ^(41)^.

Some mothers continued going to work as usual, but were not comfortable doing so. These mothers had higher odds of reporting moderate or extreme impacts of the COVID-19 pandemic. Essential workers may not have a choice to work remotely despite the high risk and anxiety surrounding COVID-19, especially if their household income depended on on their salary. A Pew research study reported that Hispanic workers had less work flexibility than other groups ^(42)^, and essential occupations linked to higher risks of infection have higher proportions of racial and ethnic minorities ^(43–46)^. US Census Bureau data indicate that women account for a disproportionate share of essential occupations in health care, education, personal care, sales, and office jobs, despite pervasive gender wage gaps ^(47)^. These results highlight racial and gender disparities that were exposed during the COVID-19 pandemic.

## Conclusions

Mothers of infants under 1 year old had increased food insecurity during the early COVID-19 pandemic (July–August 2020), although WIC use was relatively unchanged. These results suggest that food secure mothers of young children are extremely vulnerable to becoming food insecure under economic downturns, changes in childcare accessibility, and changes in work status. Our results also highlight disparities experienced by minorities and communities of color. Therefore, we recommend that vulnerable populations such as minority mothers with infants should receive specific resources and information during the early stages of socioeconomic emergencies to protect them from becoming food insecure. More research is needed to identify long-term effects of the COVID-19 pandemic on health outcomes of infants, young children, and vulnerable populations who are disproportionately affected.

## Study limitations

This study has two potential limitations. Our researchers combined Asian, Pacific Islander, American Native, and mixed/other into one group for statistical power. The survey asked participants to recall information regarding food insecurity at their present time (July–August 202) and before the COVID-19 contingency several months prior, which may lead to recall bias in baseline food insecurity levels.

## Authorship

Conceptualization, MRP; methodology, MRP; software, MRP; validation, MRP; formal analysis, ZH and MRP; investigation, ZH and MRP; resources, MRP; data curation, ZH and MRP; original draft preparation, ZH; review and editing, ZH and MRP; visualization, ZH; supervision, MRP; project administration, MRP; funding acquisition, MRP.

## Ethical standards disclosure

This study was conducted according to the guidelines laid down in the Declaration of Helsinki and all procedures involving research study participants were approved by the Institutional Review Board (or Ethics Committee) of Saint Louis University protocol code 31349, approved July 02, 2020. Written informed consent was obtained from all subjects/patients.

## Data Availability

All data produced in the present work are contained in the manuscript.

